# Evaluation of a Bayesian hierarchical pharmacokinetic-pharmacodynamic model for predicting parasitological outcomes in Phase 2 studies of new antimalarial drugs

**DOI:** 10.1101/2024.04.14.24305810

**Authors:** Meg K Tully, Saber Dini, Jennifer A Flegg, James S McCarthy, David J Price, Julie A Simpson

## Abstract

The rise of multidrug resistant malaria requires accelerated development of novel antimalarial drugs. Pharmacokinetic-pharmacodynamic (PK-PD) models relate blood antimalarial drug concentrations with the parasite-time profile to inform dosing regiments. We performed a simulation study to assess the utility of a Bayesian hierarchical mechanistic PK-PD model for predicting parasite-time profiles for a Phase 2 study of a new antimalarial drug, cipargamin.

We simulated cipargamin concentration- and malaria parasite-profiles based on a Phase 2 study of 8 volunteers who received cipargamin 7 days after inoculation with malaria parasites. The cipargamin profiles were generated from a 2-compartment PK model and parasite profiles from a previously published biologically informed PD model. One-thousand PK-PD datasets of 8 patients were simulated, following the sampling intervals of the Phase 2 study. The mechanistic PK-PD model was incorporated in a Bayesian hierarchical framework and the parameters estimated.

Population PK model parameters describing absorption, distribution and clearance were estimated with minimal bias (mean relative bias ranged from 1.7 to 8.4%). The PD model was fitted to the parasitaemia profiles in each simulated dataset using the estimated PK parameters. Posterior predictive checks demonstrate that our PK-PD model successfully captures both the pre- and post-treatment simulated PD profiles. The bias of the estimated population average PD parameters was low-moderate in magnitude.

This simulation study demonstrates the viability of our PK-PD model to predict parasitological outcomes in Phase 2 volunteer infection studies. This work will inform the dose-effect relationship of cipargamin, guiding decisions on dosing regimens to evaluate in Phase 3 trials.

## 1 Introduction

Almost 40% of the global population live in malaria endemic areas, with an estimated 249 million clinical cases in 2022, and over 608,000 deaths [1]. Following a significant fall in the global malaria burden between 2005 and 2015, the estimated number of malaria cases and deaths has begun to rise over recent years [1]. The availability of effective antimalarial drugs is key to reducing the burden of morbidity and mortality attributable to malaria.

Artemisinin-based combination therapies (ACTs), comprised of a highly potent and rapid-acting artemisinin-derivative with a longer-acting partner drug, are the current first-line treatment for *Plasmodium falciparum* malaria infection. However, partial resistance to artemisinins is now widespread across Southeast Asia [2] and more recently, has emerged *de novo* in some African countries [3, 4], South America [5] and Papua New Guinea [6]. Moreover, resistance to the partner drugs used in ACTs, such as piperaquine, has also been detected in Southeast Asia [7], resulting in treatment failures. New antimalarial drugs are urgently needed.

Drug development is a resource-heavy, expensive and time-consuming process, with only approximately 10% of drugs tested in Phase 1 trials ultimately gaining approval [8]. The journey from early phase clinical trials to Phase 3 clinical trials in patients, to then drug registration, can take many years [9]. Cipargamin is a promising candidate antimalarial drug that has transitioned from early phase studies [10] to Phase 2 clinical trials of adult patients with falciparum malaria [11, 12]. In particular, it is a rapidly acting parenteral agent with promise to replace artemisinin [13]. McCarthy and colleagues investigated the efficacy of cipargamin in a Phase 2 clinical trial [14] in 8 healthy volunteer patients who were experimentally infected with malaria and seven days later administered a low dose (10mg) of cipargamin.

These human challenge studies, also known as volunteer infection studies, involve purposeful infection of healthy volunteers in a controlled environment, and produce rich data on both parasite and drug concentrations through frequent sampling [15]. Given the ethical considerations of infecting healthy volunteers, it is imperative that the maximum information possible is obtained from these data, in order to guide selection of dosing regimens investigated for future Phase 2 and 3 studies. Statistical methods that are tailored to generating inferences from these valuable data are thus required. Pharmacokinetic-Pharmacodynamic (PK-PD) modelling is a typical framework used for such analyses. These models integrate the PK model, that describes the drug concentration over time, with a PD model that characterises the drug’s effect on the parasite population. Ideally, a PK-PD model should capture key elements of the underlying biological system, whilst remaining sufficiently simple for practical estimation and interpretation of key parameters [16].

In this study we assessed an adaptation of an existing mechanistic Bayesian hierarchical PK-PD model developed by Dini *et al.* [17], which captures the life cycle of the parasite within the red blood cell. Within a simulation-estimation framework, we investigated how precisely and accurately this model was able to recover the PK and PD parameters. The simulation study is based on data from the Phase 2 clinical study of cipargamin [14].

## 2 Results

A detailed description of the PK model, PD model, the Bayesian inference framework, and simulation study setup, including all model parameters, are provided in the Methods section. Definitions of the PK and PD model parameters are given in Tables 1 and 2 with a study overview diagram provided in Figure 1.

**Table 1:** Definitions of pharmacokinetic model parameters.

| Parameter (units) | Definition |
| --- | --- |
| $Cl$ (L/h) | Clearance rate of the drug |
| $V_c$ (L) | Central compartment volume |
| $Q$ (L/h) | Inter-compartmental clearance rate |
| $V_p$ (L) | Peripheral compartment volume |
| $k_a$ (/h) | Absorption rate |

**Table 2:** Definitions of pharmacodynamic model parameters.

| Parameter (units) | Definition |
| --- | --- |
| $i_{pl}$ (total #) | Initial Parasite Load. Total number of parasites at inoculation or model start |
| $\mu_{ipl}$ (h) | Initial mean parasite age |
| $\sigma_{ipl}$ (h) | Standard deviation of the age distribution of the initial parasite load |
| $PMF$ | Parasite multiplication factor. Number of parasites released by a ruptured schizont at the end of the life cycle |
| $E_{max}$ (% killed/h) | Maximal hourly killing rate of the drug |
| $EC_{50}$ (ng/mL) | *In vivo* drug concentration when killing rate is 50% of $E_{max}$ |
| $\gamma$ | Slope of *in vivo* drug concentration-effect curve |

**Figure 1:**
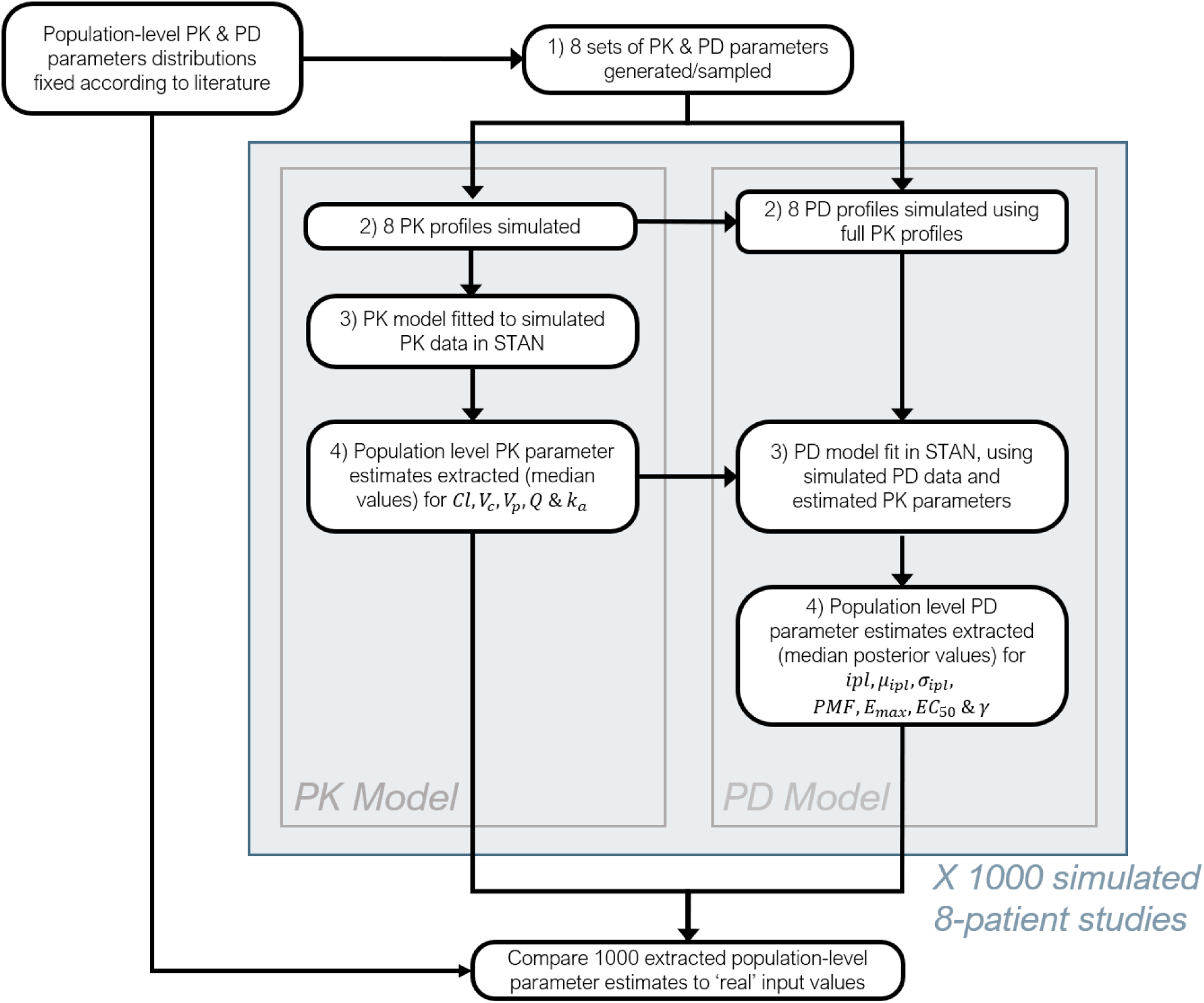
Flowchart describing the stages of the current simulation study framework.

### Pharmacokinetic Model

Cipargamin concentrations were simulated using a 2-compartment PK model with first-order absorption, based on the estimated PK parameters and between-individual variability from the analysis of the Phase 2 trial PK data [14] (Table 3). A total of 1000 simulated datasets were generated, each dataset included the PK and PD profiles of 8 patients, incorporating between- and within-individual variability. The simulated 8-patient PK datasets provided a good visual match to the trial data from McCarthy *et al.* [14] (Figure S1). The PK model was incorporated into a Bayesian hierarchical framework, and fitted to each of the 1000 simulated datasets, restricting data to the cipargamin concentrations which correspond to the sampling times of the original Phase 2 trial (1, 2, 3, 4, 6, 8, 12, 16, 24, 36, 48, 72, 96 and 120 hours post-treatment), and the posterior median estimate of each population PK parameter obtained. To evaluate how accurately this model can estimate PK parameters, we calculated the difference (absolute and relative bias) between the posterior median estimate of the population-level PK parameter, and the value used to simulate the data (*i.e.*, the ‘true’ value). Table 4 shows the ‘true’ PK parameter values used to simulate the data, the mean, 2.5- and 97.5-percentiles (herein, 95% intervals) across the 1000 posterior median estimates associated with each simulation, and the bias (absolute and relative) in these posterior median estimates. The population-level PK parameters were reliably estimated, with the magnitude of relative bias ranging from 1.7% to 8.4%, comparing the mean of the posterior median estimates to the ‘true’ value. To contextualise the bias in these estimates, we compared the PK profile created by the ‘true’ population parameters to the PK profiles generated at the 1000 posterior median parameter estimates (Figure 2). This figure demonstrates that the average PK profiles for cipargamin are captured well across all simulations.

**Figure 2:**
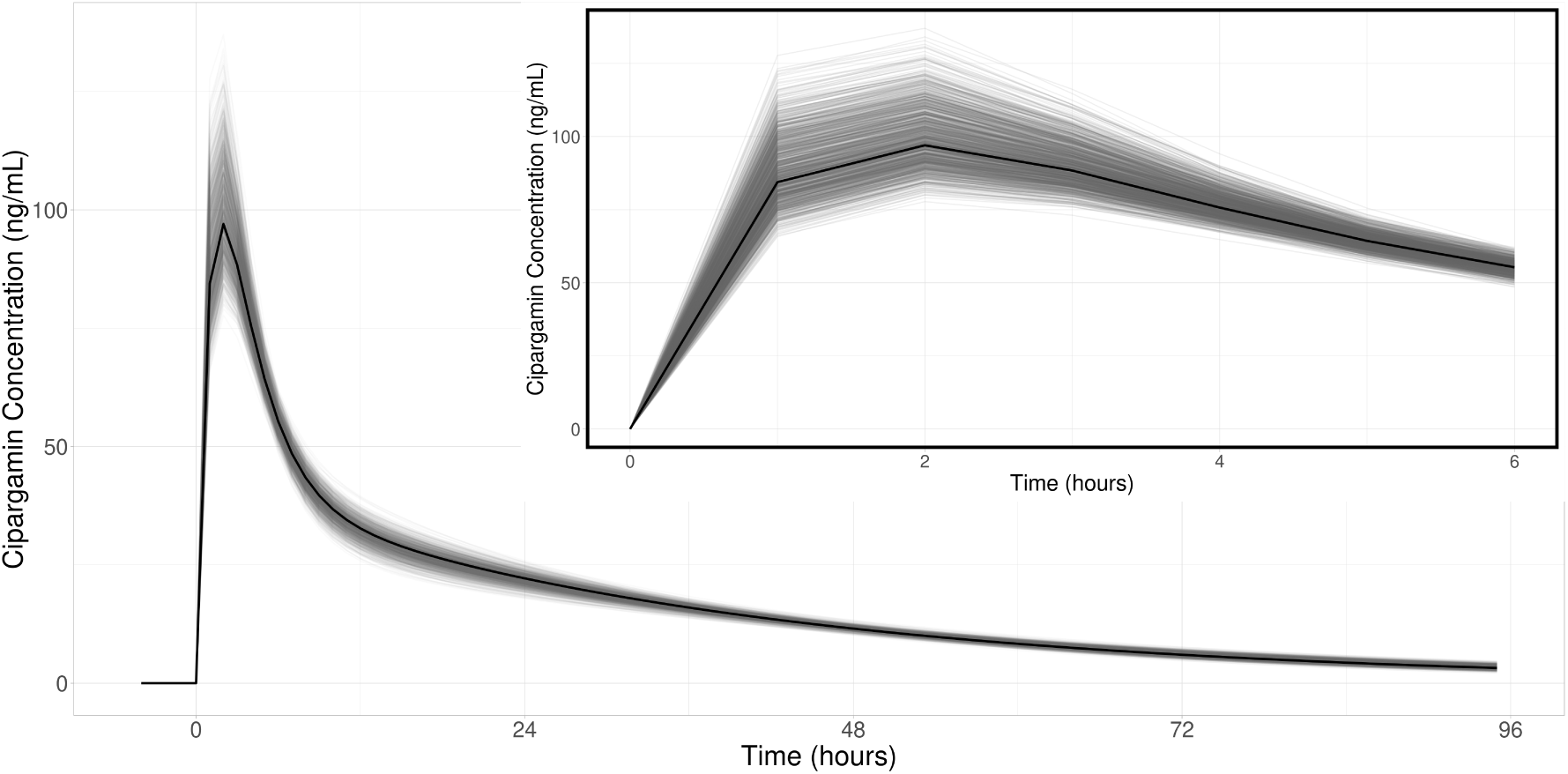
PK drug concentration profiles for a 2-compartment model produced from ‘true’ parameters values used in creating the simulations (black), compared to 1000 profiles created from each of the 1000 dataset’s mean estimated values (grey).

**Table 3:** Population parameters (***θ***), and feasible lower (***b***) and upper (***a***) bounds for each parameter in the first-order absorption two-compartment pharmacokinetic model for cipargamin.

| Parameter (units) | $\theta$ | $[\mathbf{b}, \mathbf{a}]$ |
| --- | --- | --- |
| $Cl$ (L/h) | 5.5 | [2.75, 11] |
| $V_c$ (L) | 64.4 | [32.2, 128.8] |
| $Q$ (L/h) | 12.9 | [6.45, 25.8] |
| $V_p$ (L) | 107 | [53.5, 214] |
| $k_a$ (/h) | 0.919 | [0.460, 1.838] |

**Table 4:** Mean PK parameter estimates [95% intervals] over 1000 fitted datasets, and associated bias when compared to the values used to simulate the data. Estimates are the posterior median values from a Bayesian hierarchical model.

| Parameter<br>(units) | ‘True’<br>Value | Posterior Medians | Bias |  |
| --- | --- | --- | --- | --- |
|  |  |  | Absolute | Relative (%) |
| $Cl$ (L/h) | 5.5 | 5.41 [5.08, 5.74] | -0.09 [-0.42, 0.24] | -1.64 [-7.64, 4.36] |
| $V_c$ (L) | 64.4 | 61.89 [46.52, 77.84] | -2.51 [-17.88, 13.44] | -3.90 [-27.76, 20.87] |
| $Q$ (L/h) | 12.9 | 12.36 [10.12, 14.40] | -0.54 [-2.78, 1.50] | -4.19 [-21.55, 11.63] |
| $V_p$ (L) | 107 | 111.44 [89.13, 139.44] | 4.44 [-17.87, 32.44] | 4.15 [-16.70, 30.32] |
| $k_a$ (/h) | 0.919 | 0.996 [0.83, 1.21] | 0.08 [-0.09, 0.29] | 8.38 [-9.68, 31.66] |

The population-level PK parameter least accurately estimated by the model was the absorption parameter, *k_a_*, with a mean relative bias of 8.4% [95% intervals (-9.7%, 32%)]. The PK profiles exhibit a short and sharp rise in drug concentration upon administration, during which absorption may be estimated, however the availability of only 1 to 2 observations from this period impedes the estimation of the *k_a_* parameter. When the drug concentration profiles produced from the ‘actual’ and ‘estimated’ PK parameters were compared (Figure 2), it is clear that the discrepancies between the absorption parameter values do not materially impact the cipargamin concentrations during the distribution and elimination phases.

To investigate how well this framework can recover model parameters for a single experiment, we show an example of the posterior samples compared to the ‘true’ value in Figure 3. These show that the true parameter values are contained within the range of posterior samples for each parameter, considering pairwise correlations. FigureS4 shows the posterior predictive pharmacokinetic profiles for each of the eight patients in a single experiment, again demonstrating that the posterior model fit provides an accurate characterisation of the pharmacokinetic profile.

**Figure 3:**
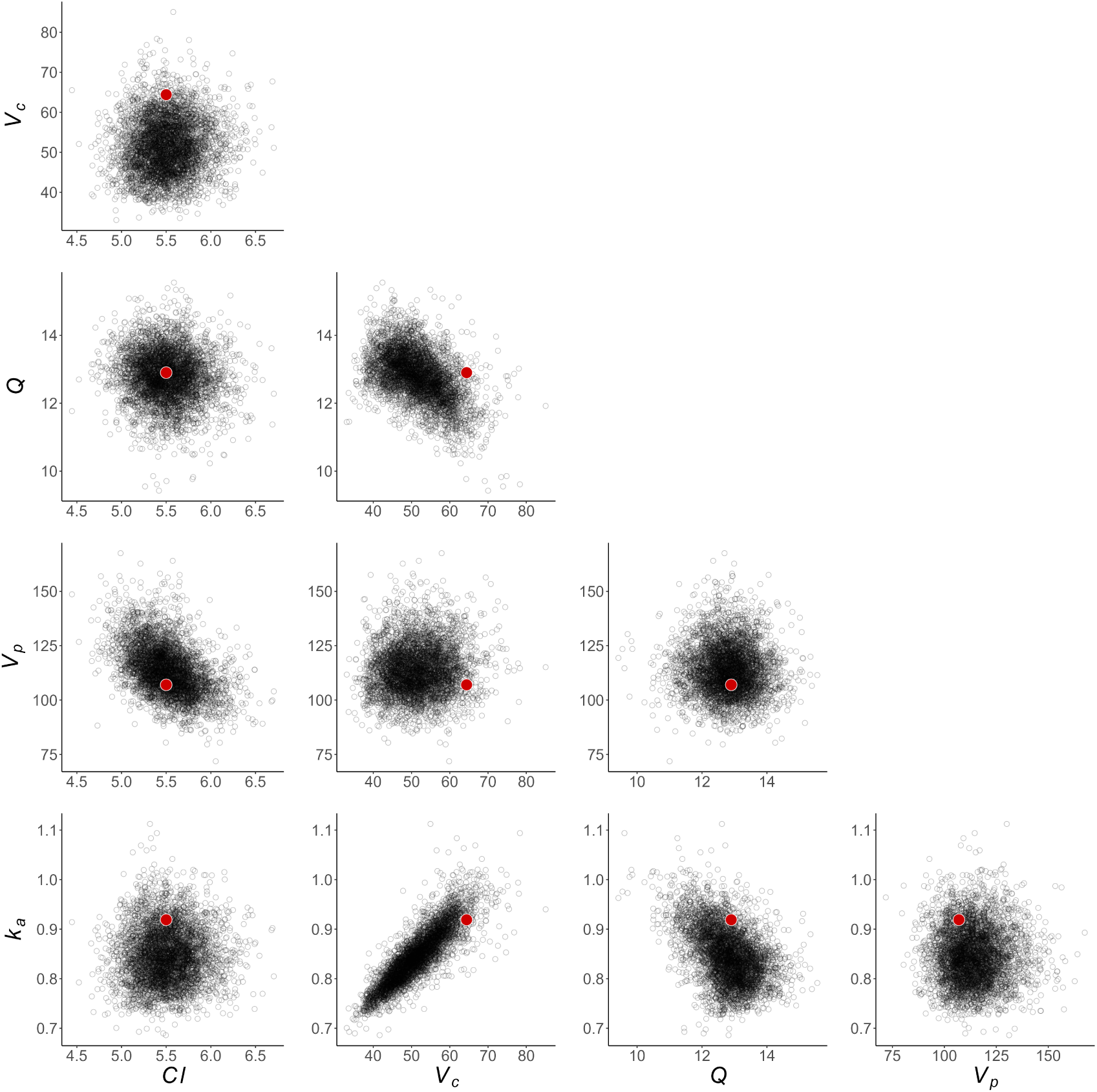
Bivariate distributions of posterior samples for population-level PK parameters, from the STAN fit of a single simulated dataset. Red dots indicate ‘true’ underlying parameter values used to simulate data.

### Pharmacodynamic Model

For each of the above simulated 1000 datasets, the 8 individual cipargamin concentration-time profiles were used to simulate 8 parasite count profiles. These parasitaemia profiles were simulated for the initial 7 days of parasite growth post-inoculation. The simulated cipargamin concentration profiles were then used to simulate drug-induced killing of the parasites over the next two days, post cipargamin administration on day 7. The PD model simulated the number of parasites aged 1 to 40 hours at each time point, and data for fitting the model was again restricted to the sampling times of the original study (72, 96, 108, 120, 132, 144, 156, 168, 172, 176, 180, 184, 192, 198, 204, 216, 228, 240, 264 and 288 hours post-innoculation). We assumed cipargamin had an immediate effect on the parasite, and that the concentration-effect relationship followed Michaelis-Menten kinetics. The PD parameter values and between-individual variability selected for generation of the PD profiles are provided in Table 5. The 1000 simulated PD datasets provided a good visual match to the parasitaemia data from McCarthy *et al.* [14] (Figure S5).

**Table 5:** Population parameters (*θ*), and feasible lower (*b*) and upper (*a*) bounds for each parameter in the pharmacodynamic model.

| Parameter (units) | $\theta$ | $[\mathbf{b}, \mathbf{a}]$ |
| --- | --- | --- |
| $i_{pl}$ (total #) | 1800 | [1500, 2100] |
| $\mu_{ipl}$ (h) | 2 | [1, 24] |
| $\sigma_{ipl}$ (h) | 3 | [1, 14] |
| $PMF$ | 13 | [5, 50] |
| $E_{max}$ (% killed/h) | 0.23 | [0.05, 1] |
| $EC_{50}$ (ng/mL) | 15.1 | [0.5, 30] |
| $\gamma$ | 5 | [0.05, 1.838] |

Table 6 shows the ‘true’ PD parameter values, the mean and 95% intervals across the 1000 posterior median estimates, and the absolute and relative bias in the posterior median estimates for each PD parameter. The magnitude of relative bias for the posterior median estimates of the seven PD parameters varied between 1% and 53%. As per the PK evaluation, we contextualised this bias by plotting a profile produced by the mean PD parameter estimates for each of the 1000 simulations, and compared these to the PD profile created by the parameters used to simulate the data (Figure 4).

**Figure 4:**
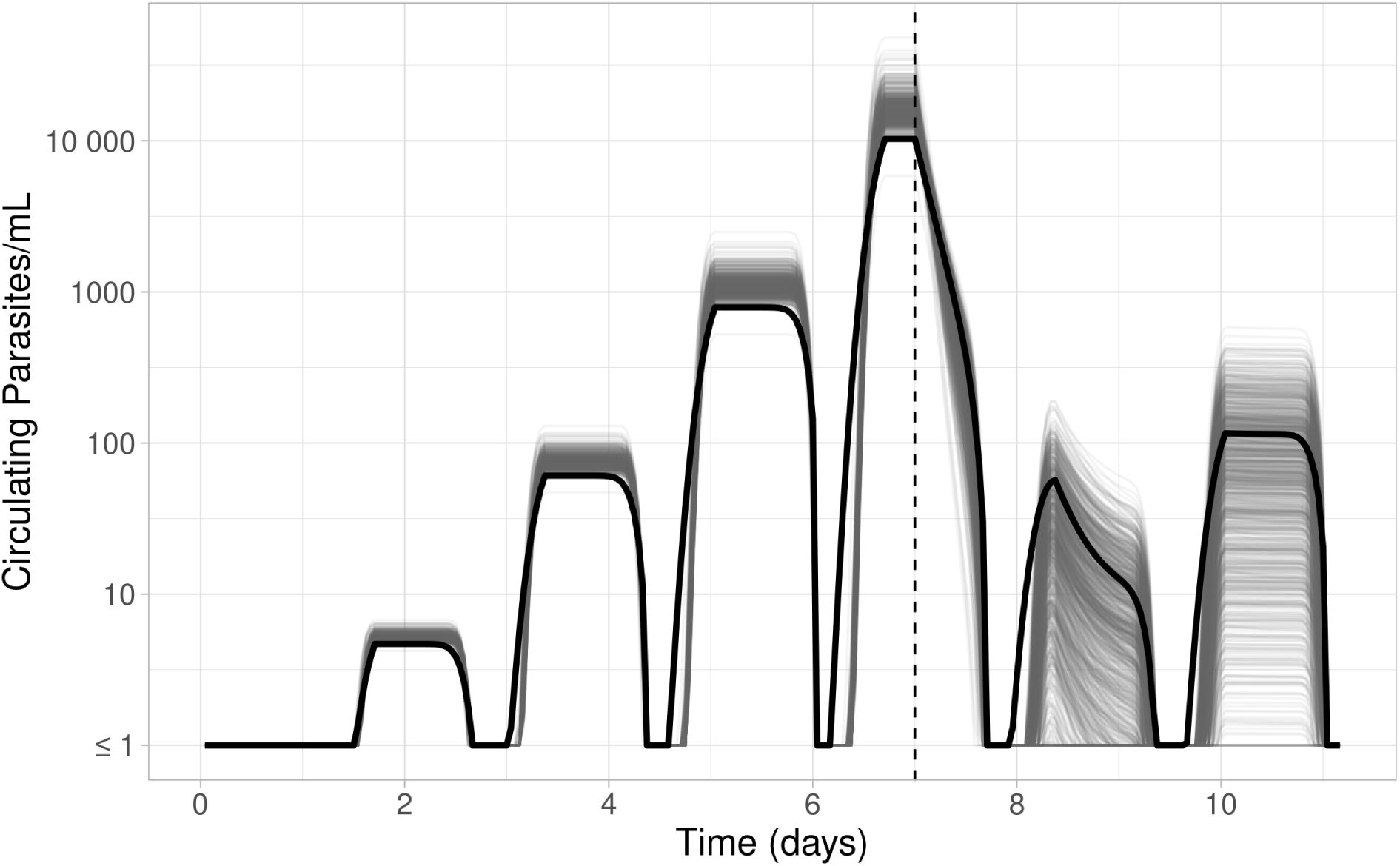
PD parasite profiles produced from ‘true’ parameters values used to create the simulations (black), compared to 1000 profiles created from each of the 1000 dataset’s mean estimated parameter values (grey). Dashed vertical line at day 7 indicates treatment.

**Table 6:** Mean PD parameter estimates [95% intervals] over 1000 fitted datasets, and associated bias when compared to the values used to simulate the data. Estimates are the posterior median values from a Bayesian hierarchical model.

| **Parameter**<br>**(units)** | **‘True’**<br>**Value** | **Posterior Medians** | **Bias** |  |
| --- | --- | --- | --- | --- |
|  |  |  | **Absolute** | **Relative (%)** |
| $i_{pl}$ ($\# \times 10^3$) | 1.8 | 1.78 [1.75, 1.82] | -0.02 [-0.051, 0.018] | -1.11 [-2.83, 1.00] |
| $\mu_{ipl}$ (h) | 2 | 2.96 [2.69, 3.44] | 0.96 [0.69, 1.44] | 48.00 [34.50, 72.00] |
| $\sigma_{ipl}$ (h) | 3 | 1.40 [1.28, 1.55] | -1.60 [-1.72, -1.45] | -53.33 [-57.33, -48.33] |
| $PMF$ | 13 | 14.55 [13.48, 16.80] | 1.55 [0.48, 3.80] | 11.92 [3.69, 29.23] |
| $E_{max}$ (% killed/h) | 0.23 | 0.29 [0.23, 0.38] | 0.06 [0.00, 0.15] | 26.09 [0.00, 65.22] |
| $EC_{50}$ (ng/mL) | 15.1 | 17.27 [13.80, 21.12] | 2.17 [-1.30, 6.02] | 14.37 [-8.61, 39.87] |
| $\gamma$ | 5 | 4.72 [2.86, 6.83] | -0.28 [-2.14, 1.83] | -5.60 [-42.80, 36.60] |

The ‘true’ mean initial parasite age (*µ_ipl_*) was 2 hours, but had a mean estimate of 2.96 hours (95% quantiles: [2.69, 3.44]). Although a seemingly large relative bias (48% [34.50%, 72.0%]), this discrepancy is less than one hour difference in parasite age. These still represent a mean age of parasites in the early ring stage of the parasite life cycle. Estimates of the standard deviation of the initial parasite age (*σ_ipl_*) are associated with a similarly large relative bias (-53.3%, [-57.3%,-48.3%]). When we compared the profiles produced by the estimated and ‘true’ values, (Figure 4) the bias in these estimates had a negligible impact on the overall parasite dynamics.

The estimated values of the PD parameters representing the maximum drug effect, *E_max_* (‘true’ value = 0.23), and the cipargamin concentration at which half of this effect is achieved, *EC*_50_ (‘true’ value = 15.1), have relatively moderate bias with mean posterior median estimates (95% quantiles) of 0.29 (0.23, 0.38) and 17.27 (13.80, 21.12) ng/ml, respectively. These estimates correspond to mean relative biases of 26.1% for *E_max_* and 14.4% for *EC*_50_. These PD parameters, together with *γ*, define the killing effect of the drug (Equation (2)). As a result, the bias in these estimates produces a noticeable discrepancy in the total number of parasites post-treatment (Figure 4).

As with the PK results, we demonstrate that this framework can recover PD model parameters (excluding the mean and spread of the initial parasite age distribution as described above) for a single experiment by presenting an example of the posterior samples compared to the ‘true’ value in Figure 5. These show that the true parameter values are well contained within the range of posterior samples for each parameter, considering pairwise correlations. Supplementary Figure S6 shows the posterior predictive PD profiles for each of the eight patients in 3 randomly selected 8-patient cohorts, again demonstrating that the posterior model fit provides an accurate characterisation of the PD profile.

**Figure 5:**
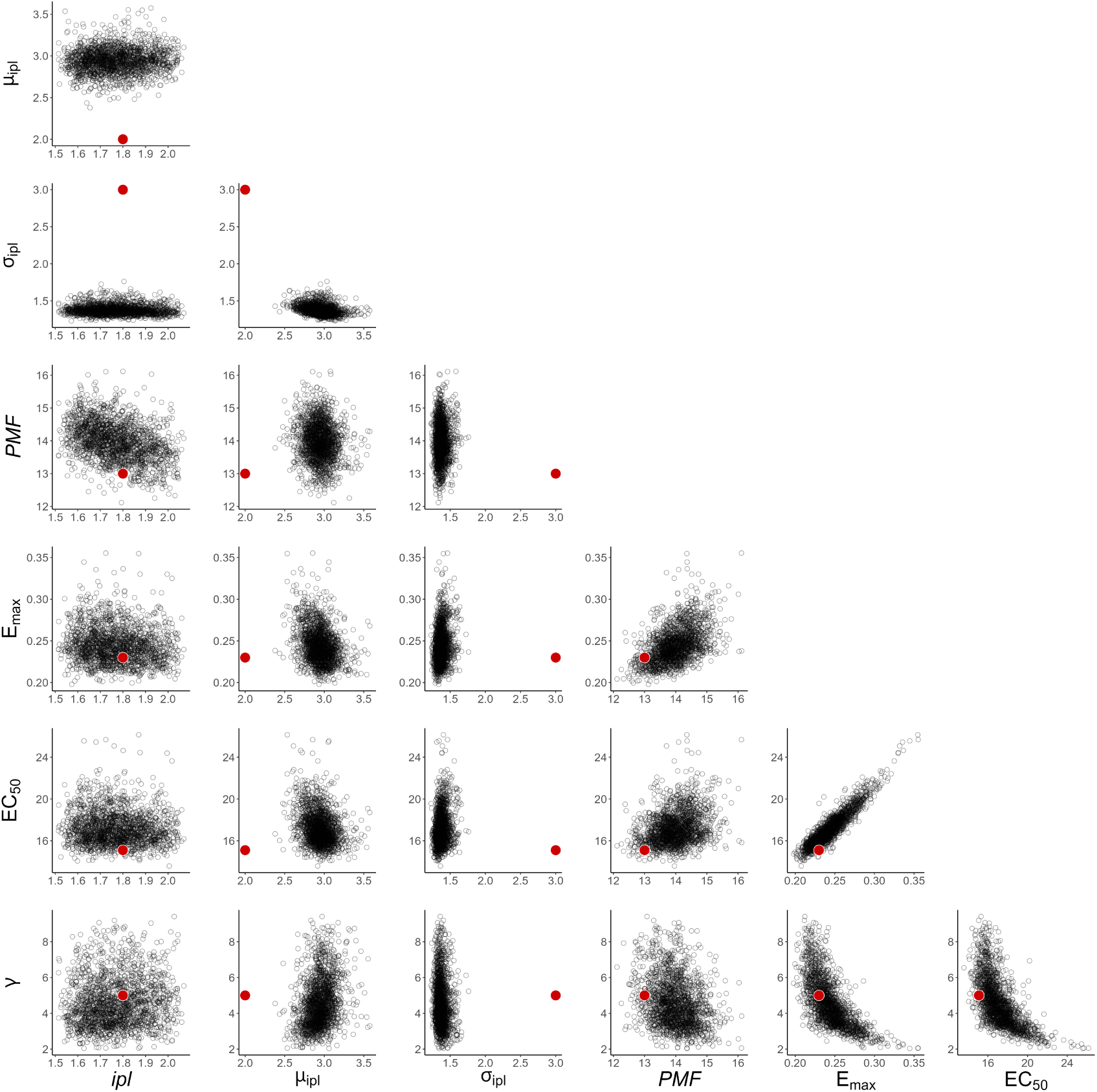
Bivariate distributions of posterior samples for population-level PD parameters, from the STAN fit of a single simulated dataset. Red dots indicate ‘true’ underlying parameter values used to simulate data.

## 3 Discussion

The results of this simulation-estimation study demonstrate that parameters of the biologically informed PK-PD model can be estimated with relatively high accuracy for Phase 2 volunteer infection studies. The PK parameters in particular were all estimated with very low bias, whereas the estimation of certain PD parameters showed comparatively less precision. In particular, the mean (*µ_ipl_*) and standard deviation (*σ_ipl_*) of the initial parasite age distribution corresponded to a relative bias of 48.0% and 53.3%. However, in absolute terms, this bias corresponds to approximately one hour in the 40-hour parasite life-cycle which does not substantially impact characterisation of parasite dynamics.

Post-treatment parasite counts are often below the limit of quantification (LOQ). This model accounts for the measurement uncertainty in those data points by averaging across the range [0, LOQ], which provides some information on the relevant parameter values, but less than contributed by points measured above the LOQ. This imperfect observation contributes to the relatively poorer estimation performance of the PD model. Generated datasets that had more post-treatment observations under the LOQ resulted in poorer modelling accuracy (Figure S7).

This form of PK-PD Bayesian hierarchical model has been previously applied to volunteer infection and patient trial datasets [17, 18]. The mechanistic form includes the hourly age of the parasite within the red blood cell for each individual, capturing the asexual reproduction cycle of the parasite and also allowing for the inclusion of the stage specific action of the antimalarial drug. Estimates of the PK-PD model parameters can be derived using different statistical methods. Maximum likelihood methods are widely used in the analysis of data from early phase antimalarial-drug trials [9, 16]. However, these methods are limited, often failing to achieve convergence unless many of the parameter values are fixed. Additionally the methods are restrictive in the incorporation of pre-existing data or knowledge. In contrast, Bayesian hierarchical methods have a number of advantages, such as incorporating prior knowledge or research, and allowing variation in both the population-level parameter values, and the correlations between the distributions from which patient-levels values are drawn.

Pharmaceutical research and development is a costly and time-consuming process [19]. Limited understanding of drug effects can result in the waste of resources though sub-optimal trial design, simultaneously diverting efforts from other candidate treatments. Therefore, careful statistical analysis and interpretation serves to not only maximise the information obtained from a study, but also has the capacity to reduce further inaccuracies; potentially limiting unnecessary risks for patients and minimising delays in antimalarial drug development — and translation into practice. In addition, further computer simulation-estimation studies can be used to determine optimal sampling designs for future Phase 2 and 3 studies (*e.g.*, [20, 21]).

Extrapolation and applicability of these simulation results is necessarily limited by the underlying assumptions of the simulation framework. This model is applied with the assumption that the underlying drug and parasite dynamics are identical to the form of the specified model. An area for further investigation would be evaluation of the impact of model misspecification on recovering biological parameters via a simulation-estimation study, whereby PK and/or PD dynamics are simulated under a different model to that used for fitting (e.g., [22]).

The model presented in this paper has been shown to reliably estimate the key population-level PK-PD parameters within the sampling framework from a Phase 2 clinical trial of cipargamin [14], using simulated data. To date, there has been no published formal assessment in a simulation study of the ability of a Bayesian hierarchical PK-PD model to reliably estimate model parameters in the context of malaria. Therefore this paper serves as an example of model performance evaluation through a simulation-estimation approach, and provides confidence in the implementation of similar mechanistic malaria models and inference framework to analyse such data. This flexible model can easily be adapted to study and evaluate emerging antimalarial compounds in the future.

## 4 Methods

Herein we describe the pharmacokinetic (PK) and pharmacodynamic (PD) models, the simulations generated from each, and the process of estimating model parameters from simulated data.

### Simulation of cipargamin pharmacokinetic profiles

This study simulated cipargamin concentrations using a standard two-compartment first-order absorption PK model with linear elimination (Text S3 S2), as described in McCarthy *et al.* [14]. The definition of each PK model parameter is given in Table 1. A hierarchical (or mixed-effects) model was used to account for the between- and within-individual variability in cipargamin concentrations.

We simulated 1000 datasets, each with PK profiles for 8 patients, following the sampling intervals from McCarthy *et al.* [14]. Table 3 contains the population PK parameters, ***θ***, and between-individual variability, ***ω***, from McCarthy *et al.* [14], and lower and upper bounds on each PK parameter. The bounds were chosen to allow a range of feasible values spanning half to double the PK estimates from McCarthy *et al.* [14].

Multiplicative error terms for individual observations were drawn from a normal distribution with a mean of 0 with variance *σ*^2^, and exponentiated. The *σ*^2^ value was generated individually for each dataset, drawn from a log-normal distribution centred at 0.1 (see Text S4 for full details)

### Pharmacodynamic Model

The PD model (presented and developed in [17, 23, 18]) is a mechanistic representation of asexual parasite replication and death during the blood stage of the infection in the presence of an anti-malarial drug, represented by a series of difference equations. Representing parasite age as an integer ranging from 1 to *T_max_*, the number of parasites that are *a* hours old at time *t*, *N* (*a, t*), is given by the number that were *a −* 1 hours old at time *t −* 1. The only unique case is the number that are 1 hour old at *t >* 0: this is given by the number of parasites that are at the end of the life-cycle (*T_max_*) at the previous time step, *N* (*T_max_, t −* 1), multiplied by the parasite multiplication factor (PMF), representing the number of new merozoites released into the blood following the asexual reproduction of the parasite at the end of its life cycle. A stage-specific killing effect of cipargamin, *E*(*a, t*), at day 7 is then applied to parasites of each age (Equation (1)). Thus, the differences equations governing the parasite distribution are:

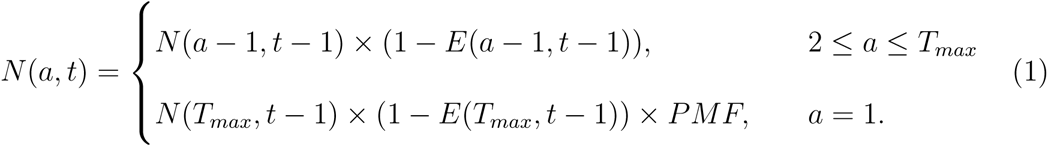

Following inoculation, the initial age-distribution, *N* (*a,* 0) is assumed to be normally distributed and discretised into hourly age groups. This distribution is defined by the number of parasites, *ipl*, and the mean, *µ_ipl_*, and standard deviation, *σ_ipl_*, of the parasite age distribution. During the growth phase, as the parasites age and replicate, the distribution shifts.

The effect of treatment on parasites of age *a* at time *t*, *E*(*a, t*), is assumed to have Michaelis-Menten kinetics, and depend on the drug concentration (*C*(*t*)) the Maximum Killing Effect (*E_max_*), the drug concentration for which 50% of that maximum killing effect is achieved (*EC*_50_), and lastly the sigmoidicity of the concentration-effect curve (*γ*):

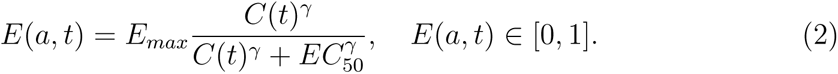

For this model, the life cycle was set to 40h in order to enable a visual match to the periodic trends of the trial data in McCarthy *et al.* [14], that were not reproducible with a 48h cycle. This is consistent with Wockner *et al.* [24], where it was found that a range of 38.3 to 39.2 hours was the reproductive cycle length most strongly supported by their data from volunteer infection studies. Although Wockner *et al.* were using a different parasite dynamic model, these estimates were based on the same strain of malaria and a population of healthy volunteers with no prior malaria infections, similar to the participants of the trial data in McCarthy *et al.* [14].

### Simulation of parasite density versus time profiles

The 1000 8-patient parasite density-time datasets were simulated using the PD model, each corresponding to one set of simulated PK data. Each profile begins with a growth-phase starting from inoculation, followed by a treatment-phase from day 7 onwards. The concentration profiles of the simulated PK data were input into the PD equation to generate the killing effect of the drug during treatment. Individual PD parameters were generated via the same approach as described for the PK parameters. That is, patient-level parameters were drawn from population-level distributions centred around ***θ***. Drug effect parameters were given by estimates from McCarthy *et al.* [14], and the parasite multiplication factor informed by [24]. Table 5 contains the population PD parameters, ***θ***, and lower and upper bounds on each parameter. Aside from PK input data, the only other factors that varied between simulations were the variance-covariance matrix and noise distribution.

### Estimation of pharmacokinetic and pharmacodynamic parameters

For each of the 1000 simulated datasets, parameters were estimated in a Bayesian framework using a Hamiltonian Monte Carlo No U-Turn Sampler in RStan v2.21.0 [25] using R version 4.1.1 [26]. For fitting the PK model to the simulated cipargamin concentrations three chains were run with 2000 iterations each and 500 discarded as warm-up. This produced 4500 posterior samples for each PK parameter, from which the posterior median was extracted as a central estimate of the posterior distribution. *R*^^^, the effective sample size (*n_eff_*), trace plots and posterior predictive interval plots were assessed to confirm that the chains had converged and were sufficiently will mixed, and that the posterior predictive distributions captured the simulated cipargamin concentration profiles accurately (Figures S2 and S3).

For Bayesian modelling of the simulated parasitaemia data, three chains were run with 1000 iterations each and 400 iterations discarded as warm-up, leaving 1800 for iterations for analysis. This was fewer than the number of iterations for each PK dataset due to a comparatively longer processing-time to evaluate the likelihood, however visual assessment of the parameter trace plots confirmed adequacy of the burn-in period and suitable convergence. The same diagnostics were evaluated as for the PK model fitting, in order to ensure chains were appropriately well-behaved, and posterior predictive distributions characterised the data (Figure S6).

### Graphical Representation

To evaluate the estimation accuracy of the PK-PD model, we compared the posterior medians to the ‘true’ underlying input values. This comparison of the posteriors medians (mean [95% intervals]) is presented in Tables 4 (for PK parameters) and 6 (PD parameters). Additionally, we plotted the hypothetical profiles that would be produced by each set of posteriors median parameter values. These profiles are presented in Figures 2 and 4 alongside the profile that would be produced by the true population values (i.e. centres of the population parameter distributions).

Figures 3 and 5 present the full distribution of all posterior samples from the STAN fit of a randomly selected single dataset.

All statistical computing code for the simulation and estimation steps is available at https://github.com/M-Tully/pkpd_cipargamin_model.

## Data Availability

No new datasets are presented in this research. Code to perform the analyses is available at https://github.com/M-Tully/pkpd_model_cip

