## Supplemental Materials for "Evaluation of a Bayesian hierarchical pharmacokinetic-pharmacodynamic model for predicting parasitological outcomes in Phase 2 studies of new antimalarial drugs"




CONFLICT OF INTEREST

All authors declared no competing interests for this work.

FUNDING

This work was supported by the Australian National Health and Medical Research Council (NHMRC) Leadership Investigator Grants (#1196068 and #2016396) to JAS and JSM, the Australian Centre for Research Excellence in Malaria Elimination (#2024622) and a NHMRC Synergy Grant (#2018654).

### Contents

**S1 Pharmacokinetic Diagnostic Checks** **3**

**S2 Pharmacodynamic Diagnostic Checks** **7**

**S3 Pharmacokinetic Model** **10**

**S4 Hierarchical Simulations** **12**

### S1 Pharmacokinetic Diagnostic Checks

Visual comparison of simulations was performed to check for compatibility between the simulated data sets and Phase 2 trial data of volunteers [1]. An example of two simulated data sets are shown in Figure S1. Recall, for each simulated data

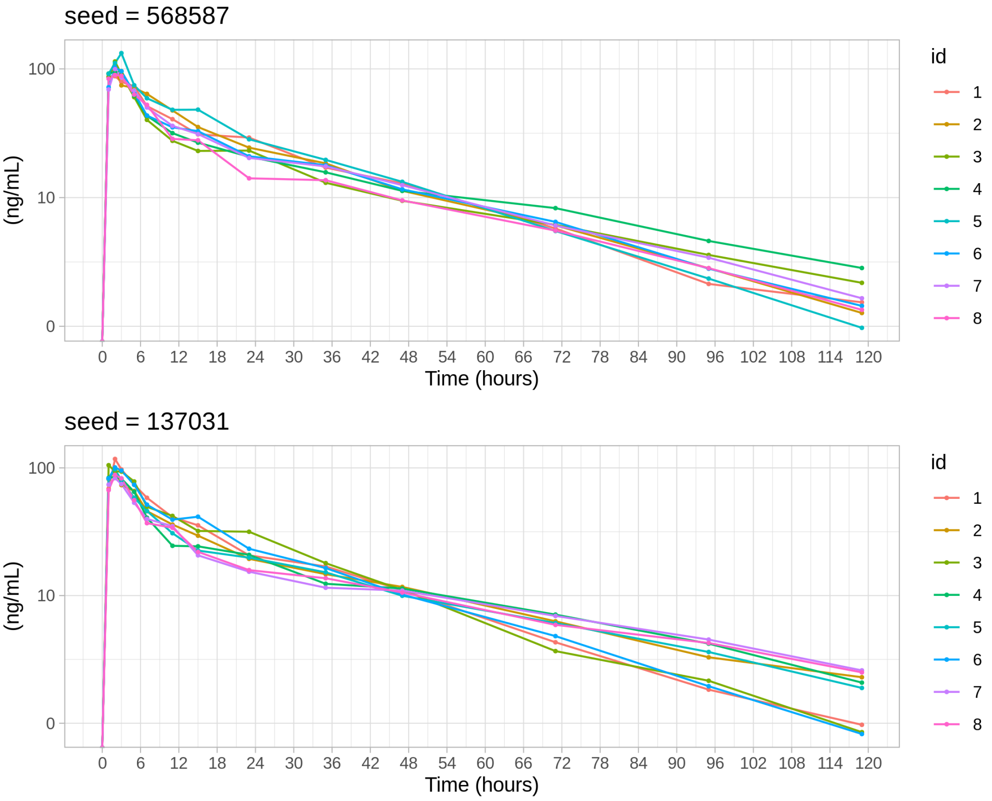


Figure S1: Examples of 2 of the 1000 simulated data sets, each comprising 8 individual patient PK drug profiles for concentration of cipargamin (ng/mL) over time (hours).

set consisting of 8 participants, three chains were run with 2000 iterations each, and 500 samples discarded as warm-up. To asses model convergence, trace plots were produced (Figure S2) and $\hat{R}$ and $n_{eff}$ statistics reviewed [2]. For the population and individual-level parameters in the example data set featured in Figure S2, the mean effective sample size was 2354 and the minimum was 352. The $\hat{R}$ values diverged from 1 by less than 0.005. These characteristics are consistent with well-mixed and converged chains [2].

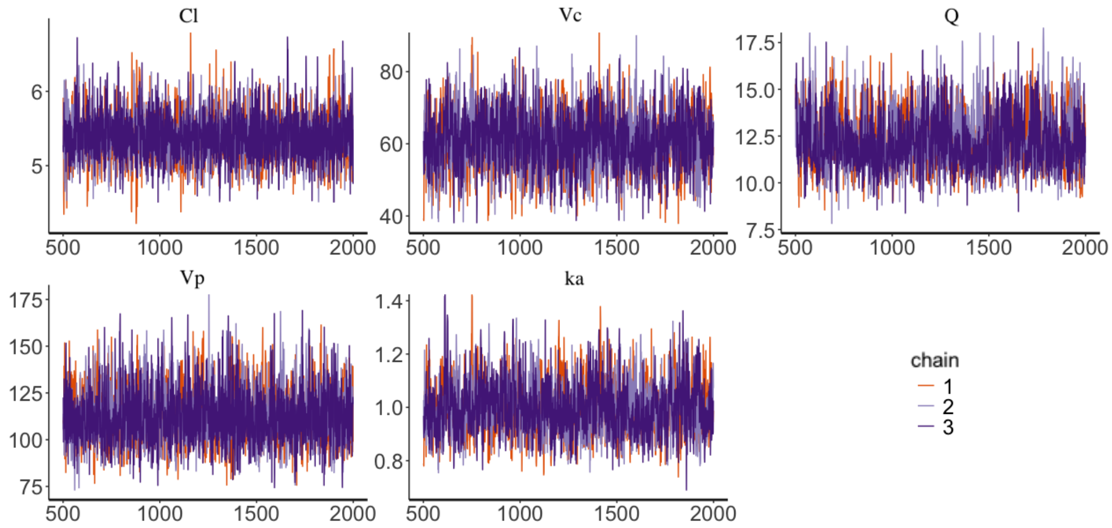


Figure S2: Trace plots from 3 independent chains for 5 population-level PK parameters, from an MCMC sampler run in Stan to an example 8-patient simulated PK data set. The first 500 of the 2000 simulations are discarded from each chain as warm-up.

When combined, the 3 chains produce 4500 iterations for each parameter, from which a single median values is calculated as the final estimate. This process is repeated over the 1000 datasets. The density plots in Figure S3 illustrate the distribution of the 1000 posterior median estimates for the 5 population parameters, one from each simulated dataset. The 95% intervals of these distributions are shown as blue shaded regions.

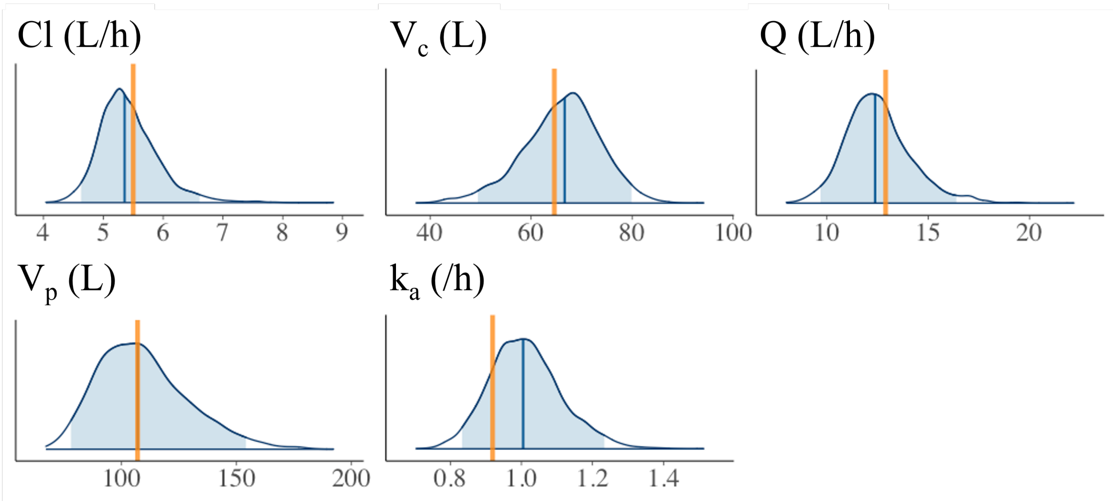


Figure S3: Posterior density plots for 5 population-level PK parameters corresponding to a single example 8-patient simulated PK data set. The shaded blue regions represent the 2.5% to 97.5% quantiles, with the posterior median shown as a vertical blue line. The orange line is the parameter value used to simulate the data.

Further evaluation of the model performance was performed via inspection of the 95% posterior predictive distributions for each of the 8 patients in a ranomdly selected sample of datasets, one of which is presented in Figure S4. The model fits appear sensible, with only points near the peak falling outside the credible intervals for some patients.

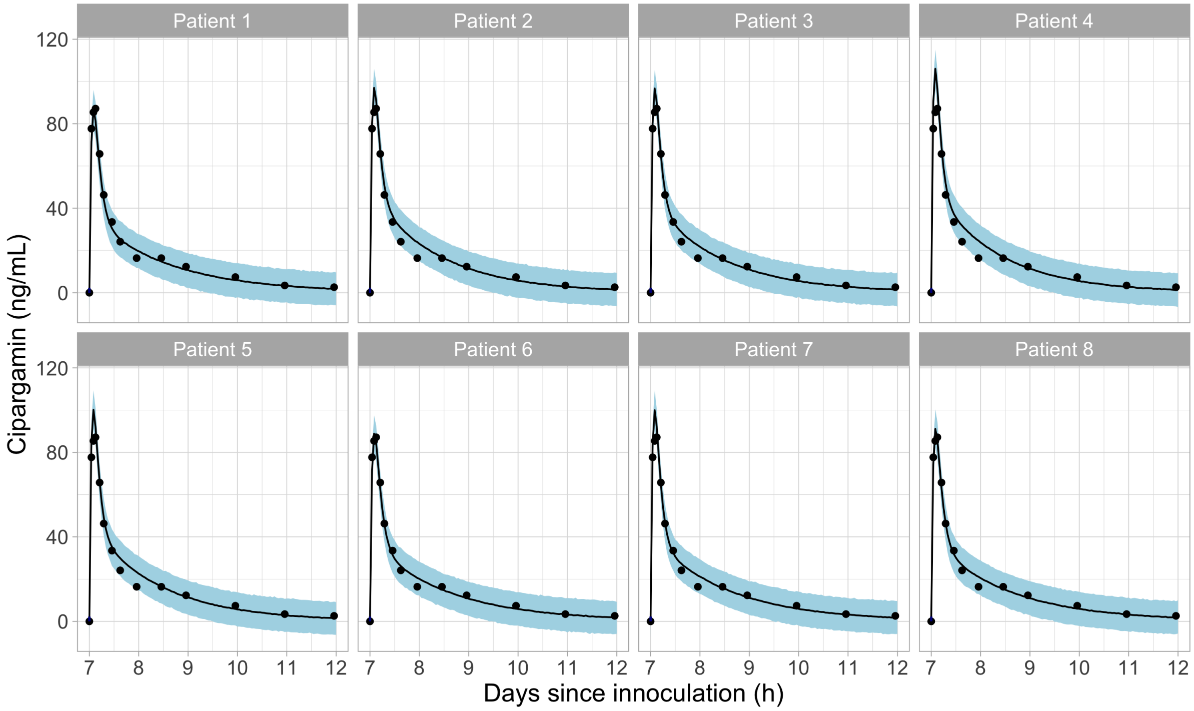


Figure S4: A single simulated dataset of 8 individual patients, with 95% posterior predictive intervals for concentration of cipargamin (ng/mL) over time (h) shaded in blue. The black line is the median posterior profile and the simulated data points are the black circles.

### S2 Pharmacodynamic Diagnostic Checks

As for the pharmacokinetic model, we visually inspected the simulated parasitaemia time profiles to confirm a good match between the simulated (Figure S5) and real Phase 2 trial data of volunteers [1]. We note a small difference in the post-treatment minimum parasite count, which occurs 8.5 days after inoculation in the study data, and at day 8 in the simulated data.

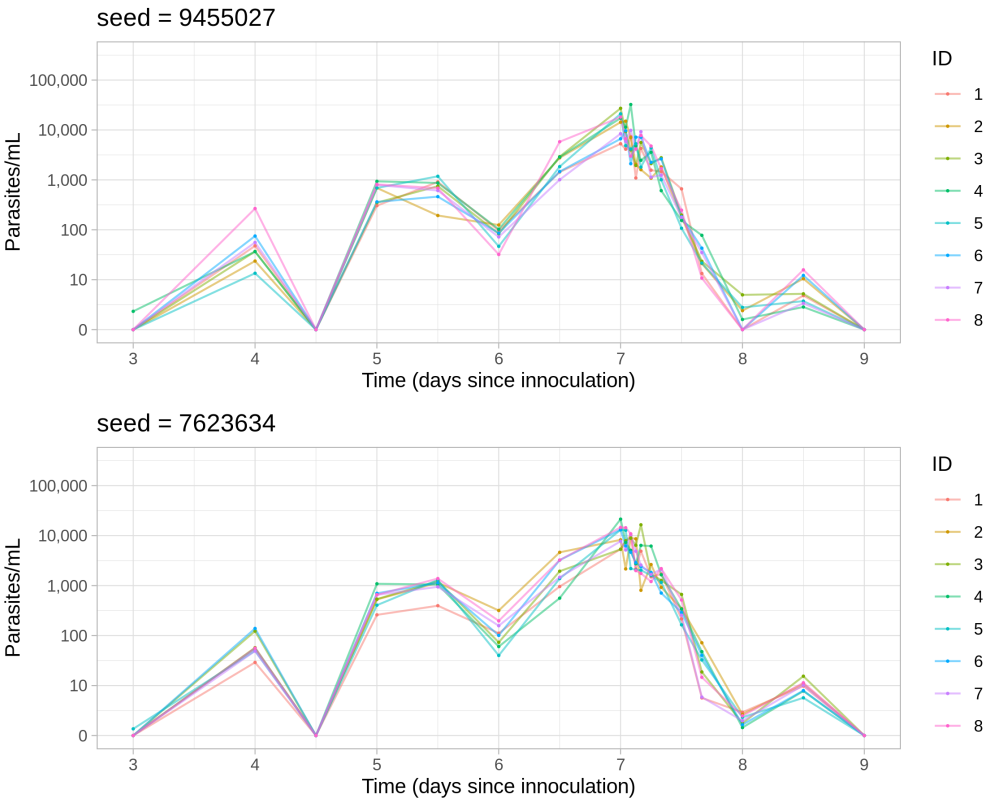


Figure S5: Two example simulated data sets, each comprising 8 individual patient PD profiles for concentration of parasites (ng/mL) over time (hours).

The 8 patient posterior predictive intervals were plotted for a random sample of 3 data sets (Figure S6) which demonstrated that the model was able to consistently capture the data well.

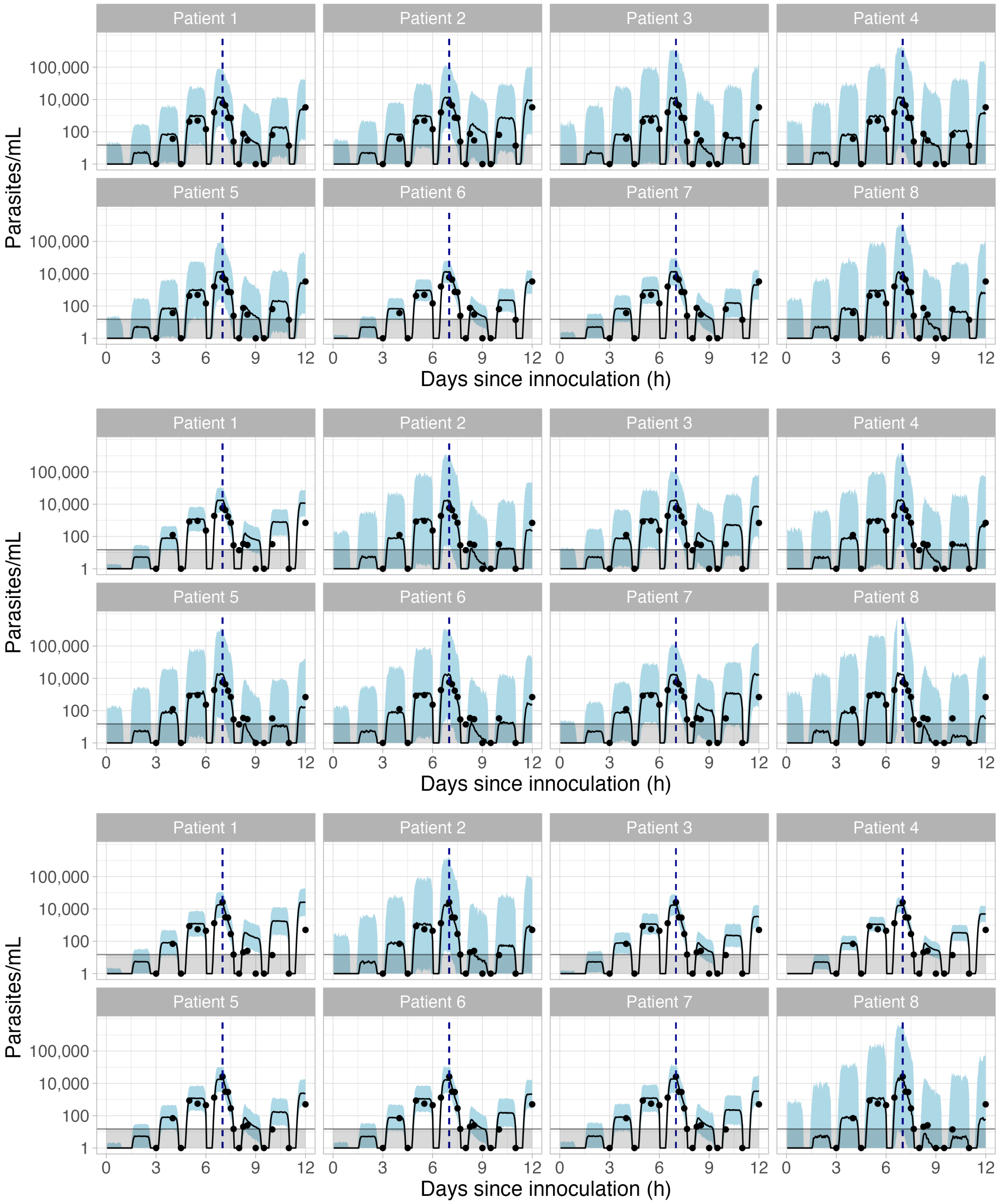


Figure S6: 95% posterior predictive distributions (blue shading) of parasite time profiles (days since inoculation) for each of 3 randomly selected simulations consisting of 8 individual patients. The black line is the median posterior profile and the simulated data points (with noise removed) are the black circles. The grey shaded area represents the region of the y-axis below the LLOQ (50 parasites/mL) and the vertical dashed line is the time the drug is administered (7 days post-inoculation).

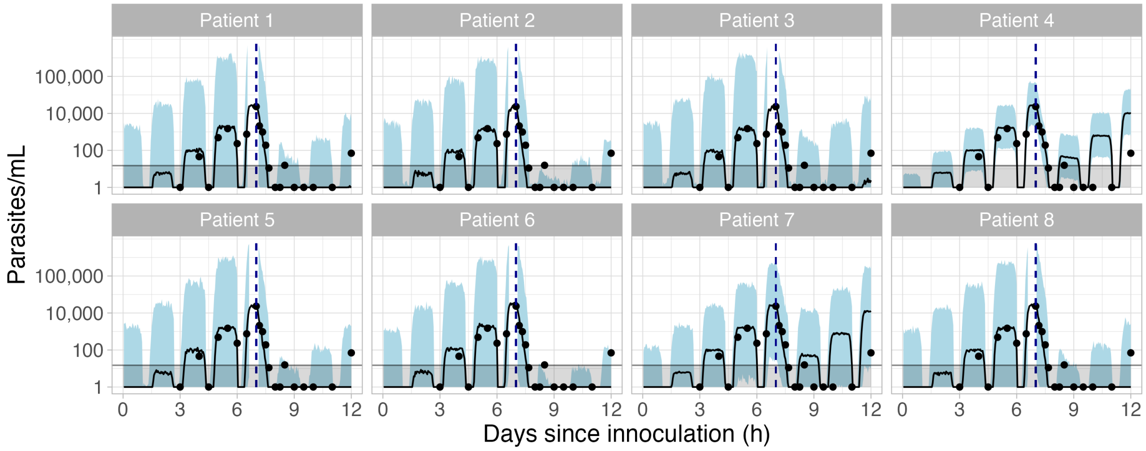


Figure S7: An example of a dataset where the post-treatment parasitaemia is underestimated.

### S3 Pharmacokinetic Model

The 2-compartment $1^{st}$ order absorption PK model with linear elimination has the following structure:

$$C(t) = D(Ae^{-\alpha t^*} + Be^{-\beta t^*} + Ce^{-t^*} - (A + B + C)e^{-k_a t^*}),$$

where

- $C(t)$ = Concentration at time $t$ (mg/L)
- $D$ = Dose (mg)
- $t$ = Time (h)
- $t_{lag}$ = Absorption lag time (h)
- $t_{dose}$ = Time of dose (h)
- $Cl$ = Elimination clearance (L/h)
- $V_c$ = Central compartment volume (L)
- $Q$ = Inter-compartmental clearance rate (L/h)
- $V_p$ = Peripheral compartment volume (L)
- $k_a$ = Absorption rate constant ($\text{h}^{-1}$)

and

- $\alpha = \left(\frac{Cl}{V_c}\frac{Q}{V_p}\right)/\beta$
- $\beta = \frac{1}{2}\left(\frac{Cl}{V_c} + \frac{Q}{V_c} + \frac{Q}{V_p} - \sqrt{(\frac{Cl}{V_c} + \frac{Q}{V_c} + \frac{Q}{V_p})^2 - 4\frac{Cl}{V_c}\frac{Q}{V_p}}\right)$
- $A = \frac{k_a}{V_c}\frac{Q/V_p - \alpha}{(k_a - \alpha)(\beta - \alpha)}$
- $B = \frac{k_a}{V_c}\frac{Q/V_p - \beta}{(k_a - \beta)(\alpha - \beta)}$

- $C = \frac{-(A(k_a-\alpha)+B(k_a-\beta))}{k_a}$
- $t^* = t - t_{lag} - t_{dose}$

The trial data from McCarthy *et al.* [1] shows a drop in parasitaemia immediately after drug administration. Therefore, we assumed cipargamin had a direct impact on the parasites, and no effect-delay parameters (i.e., $k_{e0}$, $t_{lag}$) were included in the model.

### S4 Hierarchical Simulations

Each of the 8 patient's individual PK parameters, $\boldsymbol{\theta}_i$, $(Cl, V_c, Q, V_p, K_a)$, were drawn from distributions centred at the population-level parameter estimates ($\boldsymbol{\theta}$, Table 3) via the following procedure. We represented individual PK parameters, $\boldsymbol{\theta_i}$, $i = 1, \ldots, 8$, using a logistic transformation with bounds to ensure biologically plausible values,

$$
\begin{aligned}
\boldsymbol{\phi}_i &= \log\left(\frac{\boldsymbol{\theta_i} - \boldsymbol{a}}{\boldsymbol{b} - \boldsymbol{\theta_i}}\right) \qquad \text{(Fraction is } \textbf{element-wise}\text{)} \\
&= \log\left(\frac{\boldsymbol{\theta} - \boldsymbol{a}}{\boldsymbol{b} - \boldsymbol{\theta}}\right) + \boldsymbol{\eta}_i \\
&= \boldsymbol{\phi} + \boldsymbol{\eta_i},
\end{aligned}
$$

where $\boldsymbol{\phi_i}$ are logistic-transformed population average PK parameters, vectors $\boldsymbol{a}$ and $\boldsymbol{b}$ are vectors containing the upper and lower bounds (respectively) for each parameter, and $\boldsymbol{\eta}_i$ are multivariate normal-distributed *variations* of each individuals parameter values from the logistic-transformed population averages $\boldsymbol{\phi}$. The individual deviations from these averages followed a multivariate normal distribution with mean $\mathbf{0}$ and covariance matrix $\boldsymbol{\Sigma}$. That is,

$$\boldsymbol{\eta}_i \sim MVN(\mathbf{0}, \boldsymbol{\Sigma}).$$

The variance matrix $\boldsymbol{\Sigma}$ was set to the between-individual standard deviation for each PK parameter, $\boldsymbol{\omega}$, estimated in the target trial [1], and correlation between the PK parameters was sampled from a standard uniform distribution separately for each of the 1000 simulated datasets, such that:

$$\boldsymbol{\Sigma} = \text{diag}(\boldsymbol{\omega})\mathbf{R}\text{diag}(\boldsymbol{\omega}), \quad \mathbf{R} = \mathbf{L}\mathbf{L}^T,$$

where $\mathbf{L}$ is a lower-diagonal matrix with entries $a_i$ distributed uniformly between 0 and 1:

$$\text{diag}(\boldsymbol{\omega}) = \begin{bmatrix} 0.325 & 0 & 0 & 0 & 0 \\ 0 & 0.227 & 0 & 0 & 0 \\ 0 & 0 & 0.1 & 0 & 0 \\ 0 & 0 & 0 & 1 & 0 \\ 0 & 0 & 0 & 0 & 0.1 \end{bmatrix} \quad \mathbf{L} = \begin{bmatrix} a_1 & 0 & 0 & 0 & 0 \\ a_2 & a_3 & 0 & 0 & 0 \\ a_4 & a_5 & a_6 & 0 & 0 \\ a_7 & a_8 & a_9 & a_{10} & 0 \\ a_{11} & a_{12} & a_{13} & a_{14} & a_{15} \end{bmatrix}, a_i \sim U(0,1).$$

Finally as described in the main text, multiplicative error terms for individual observations were drawn from a normal distribution with a mean of 0 with variance $\sigma^2$, and exponentiated. The $\sigma^2$ value was generated individually for each dataset, drawn from a log-normal distribution centred at 0.1.

The PD model follows an essentially identical structure for the 7 key parameters, $\boldsymbol{\theta}_i$; $(ipl, \mu_{ipl}, \sigma_{ipl}, PMF, E_{max}, EC_{50}, \gamma)$, Table 5. The input values for the diagonal matrix $\boldsymbol{\omega} = \begin{bmatrix} 0.2 & 0.2 & 0.2 & 0.0242 & 0.2 & 0.2 & 0.2 \end{bmatrix}$ were based upon the between-subject variances estimated from the analysis of the trial data in McCarthy *et al.* (2021) [1], but again these values are chosen to be a base for the random variance-covariance matrix.
